# Monogenic epilepsies exhibit distinct sleep endophenotypes

**DOI:** 10.64898/2026.06.26.26356702

**Authors:** Katharina S. Bochtler, Alexander I. Batterman, Hyun Yong Koh, Riley Kessler, Christine Esparza, Joy Shon, Michael C. Kaufman, Ingo Helbig, Vishnu Anand Cuddapah

**Affiliations:** Division of Neurology and Developmental Neuroscience, Department of Pediatrics, Baylor College of Medicine, Houston, TX, 77030, USA; Jan and Dan Duncan Neurological Research Institute and Gordon and Mary Cain Research Foundation Laboratories, Texas Children’s Hospital, Houston, TX, 77030, USA; Department of Biomedical Sciences, Cooper Medical School of Rowan University, Camden, NJ, 08103, USA; The Epilepsy NeuroGenetics Initiative (ENGIN), Children’s Hospital of Philadelphia, Philadelphia, PA, 19104, USA; Department of Biomedical and Health Informatics (DBHi), Children’s Hospital of Philadelphia, Philadelphia, PA, 19104, USA; Department of Neurology, Perelman School of Medicine, University of Pennsylvania, Philadelphia, PA, 19104, USA; Division of Neurology, Department of Pediatrics, Children’s Hospital of Philadelphia, Philadelphia, PA, 19104, USA; Liberty University College of Osteopathic Medicine, Lynchburg, VA, 24502, USA

**Keywords:** Genotype-phenotype correlation, EEG, Bayesian analysis, Neurodevelopmental disorders, Refractory epilepsy

## Abstract

Monogenic epilepsies are 1.6 times more likely to be treatment-resistant compared to other epilepsies, emphasizing the need for additional therapeutic strategies. Sleep dysfunction beyond sleep-related breathing disorders is common yet insufficiently characterized and treated in monogenic epilepsies. We therefore sought to study sleep phenotypes across these epilepsies, examine associations with seizure severity, and assess the diagnostic rate of sleep disorders.

From 2,519 individuals enrolled in the Epilepsy Genetics Research Project at Children’s Hospital of Philadelphia, we identified the monogenic epilepsies most frequently associated with sleep-related diagnoses, yielding 252 individuals across nine genetic diagnoses (*STXBP1*, *n =* 79; *SCN1A*, *n =* 57; *SCN2A*, *n =* 34; *KCNQ2*, *n =* 21; *SLC6A1*, *n =* 14; *SYNGAP1*, *n =* 13; *WDR45*, *n =* 13; *KCNT1*, *n =* 11; *PCDH19*, *n =* 10).

Monogenic epilepsies exhibited distinct sleep endophenotypes, including insomnia, parasomnia, and sleep-related movement disorders in *SCN1A*-related disorders; frequent epileptiform discharges in sleep with insomnia symptoms in *SCN2A*-related disorders; sleep dysfunction restricted to the developmental and epileptic encephalopathy subtype in *KCNQ2*-related disorders; and insomnia without nocturnal seizure involvement in *SYNGAP1*-related disorders. Formal sleep diagnoses were present in only 25% of individuals (63/252), yet 58% (145/252) reported sleep difficulties, suggesting substantial underdiagnosis. Persistent seizures were associated with higher odds of sleep disorder diagnoses (*OR* 2.87, *95% CrI* 1.57–5.36), disrupted sleep architecture (*OR* 2.06, *95% CrI* 1.08–4.16), nocturnal seizures (*OR* 4.47, *95% CrI* 2.50–8.28), hypersomnolence (*OR* 2.38, *95% CrI* 1.27–4.58) and insomnia (*OR* 1.80, *95% CrI* 1.06–3.05). Neuropsychiatric comorbidities were independently associated with sleep burden after adjustment for seizure severity (*OR* 2.49, *95% CrI* 1.40–4.49).

We find that monogenic epilepsies exhibit distinct, gene-specific sleep endophenotypes that are underdiagnosed. Treating sleep difficulties beyond obstructive sleep apnoea may improve seizure control and developmental outcomes, highlighting the need for timely diagnosis of co-occurring sleep disorders.

## Introduction

Epilepsy is one of the most common neurological disorders, affecting approximately 65 million individuals globally, with a lifetime prevalence of 7.6 in every 1,000 and a bimodal distribution with increased incidence in the young (<18 years) and old (>50 years).^1,2^ At least 30-50% of childhood epilepsies have a genetic aetiology, and genetic epilepsies that manifest before the age of three years are estimated to present in 1:2000 live births.^3,4^ Notably, genetic epilepsies are 1.6 times more likely to be treatment-resistant compared to epilepsies without a known aetiology, underscoring the need to identify additional strategies to improve outcomes in this population.^5,6^

As diagnosis of monogenic epilepsies grows with increased accessibility of molecular testing, efforts to identify personalized treatments have increased. Well-recognized examples of this include the use of the ketogenic diet for treatment of epilepsy caused by GLUT1 (*SLC2A1*) deficiency syndrome or high-dose sodium channel blockers for *SCN2A*-related epilepsy. However, despite ongoing efforts to identify aetiology-specific treatments, many patients remain medically refractory. Comorbid conditions represent an underappreciated but potentially modifiable contributor to this refractory burden. A recent meta-analysis demonstrated that children with epilepsy and neuropsychiatric disorders have higher rates of drug resistance^7^ and treating comorbidities, including ADHD^8^, depressive symptoms,^9,10^ and anxiety improves seizure control.^11^ Sleep dysfunction represents a similarly actionable comorbidity: Highly prevalent, frequently undertreated, and mechanistically linked to seizure burden, yet it remains systematically uncharacterized in genetically defined epilepsies.^12–16^

One-half to one-third of people with epilepsy have at least one sleep-related complaint, with insomnia, daytime sleepiness and hypersomnia being among the most common.^17^ In adults with epilepsy, seizure risk was 2.8 times higher with self-reported poor sleep quality and 1.9 times higher with reported insomnia.^17–19^ Approximately one-third of individuals with epilepsy have obstructive sleep apnoea (OSA),^20^ and its treatment improves both sleep quality and seizure control.^21^ Melatonin supplementation decreases seizure severity and improves sleep quality and quantity in individuals with epilepsy.^22,23^ Thus, sleep dysfunction is highly prevalent in individuals with epilepsy, and treatment of sleep dysfunction can improve seizure management.

The relationship between sleep and seizures is thought to be bidirectional: Poor sleep may worsen seizure control, while seizure burden itself may disrupt sleep architecture, creating a cycle that may be amenable to intervention at multiple points.^24,25^ Despite this, sleep disorders in epilepsy are thought to be substantially underdiagnosed in clinical practice, with formal diagnoses capturing only a fraction of the true symptom burden.^26^ While these relationships are increasingly recognized in general epilepsy populations^25,27^, whether distinct sleep endophenotypes exist across monogenic epilepsies and the extent to which sleep difficulties go undiagnosed remain largely uncharacterized.

Given that improvement of sleep quantity and quality decreases seizure burden, we sought to characterize sleep phenotypes in monogenic epilepsies. We identified the most common monogenic epilepsies with comorbid sleep dysfunction diagnosed at Children’s Hospital of Philadelphia: *STXBP1*-, *SCN1A*-, *SCN2A*-, *KCNQ2*-, *SLC6A1*-, *KCNT1*-, *SYNGAP1*-, *WDR45*- and *PCDH19*-related neurodevelopmental disorders. We found distinct sleep endophenotypes in monogenic epilepsies and highlight the importance of early detection and treatment of sleep dysfunction to improve seizure control.

## Materials and methods

### Subject recruitment and data retrieval

We identified 2,519 unrelated individuals with neurodevelopmental disorders seen at the Children’s Hospital of Philadelphia (CHOP) enrolled in the Epilepsy Genetics Research Project (EGRP; Figure 1A). Legal guardians provided informed consent for every participant, and all study procedures were conducted under protocols approved by the Children’s Hospital of Philadelphia Institutional Review Board. Patient encounters occurred from October 2015 to January 2023. The EGRP cohort was filtered in the following stepwise manner: (1) Having a sleep-related diagnosis (including insomnia, parasomnia, hypersomnia, or circadian change); and (2) Identification of a genetic aetiology for their neurodevelopmental disorder. Individuals with a genetic diagnosis with a frequency ≥ 3 occurrences (encompassing nine different genes) were included in the study (“sleep diagnosis” group). Diagnoses related to sleep-related breathing disorders (including OSA) and drug-induced sleep disorders were excluded from this round of data filtering. Since treatment of OSA has already been previously well-characterized to improve seizure control,^18^ we focused our analyses on other sleep-related diagnoses. An initial analysis in March 2022 was followed by a subsequent analysis in January 2023 to identify children with one of the previously mentioned nine monogenic aetiologies but without a sleep diagnosis. These individuals were identified through a similar process of filtering of the EGRP cohort (allocated to the “no sleep diagnosis” group). Upon manual review, two participants were appropriately moved from the “no sleep diagnosis” to the “sleep diagnosis” group. Sample size and demographic information for each genetic diagnosis are reported in Table 1.

**Figure 1.**
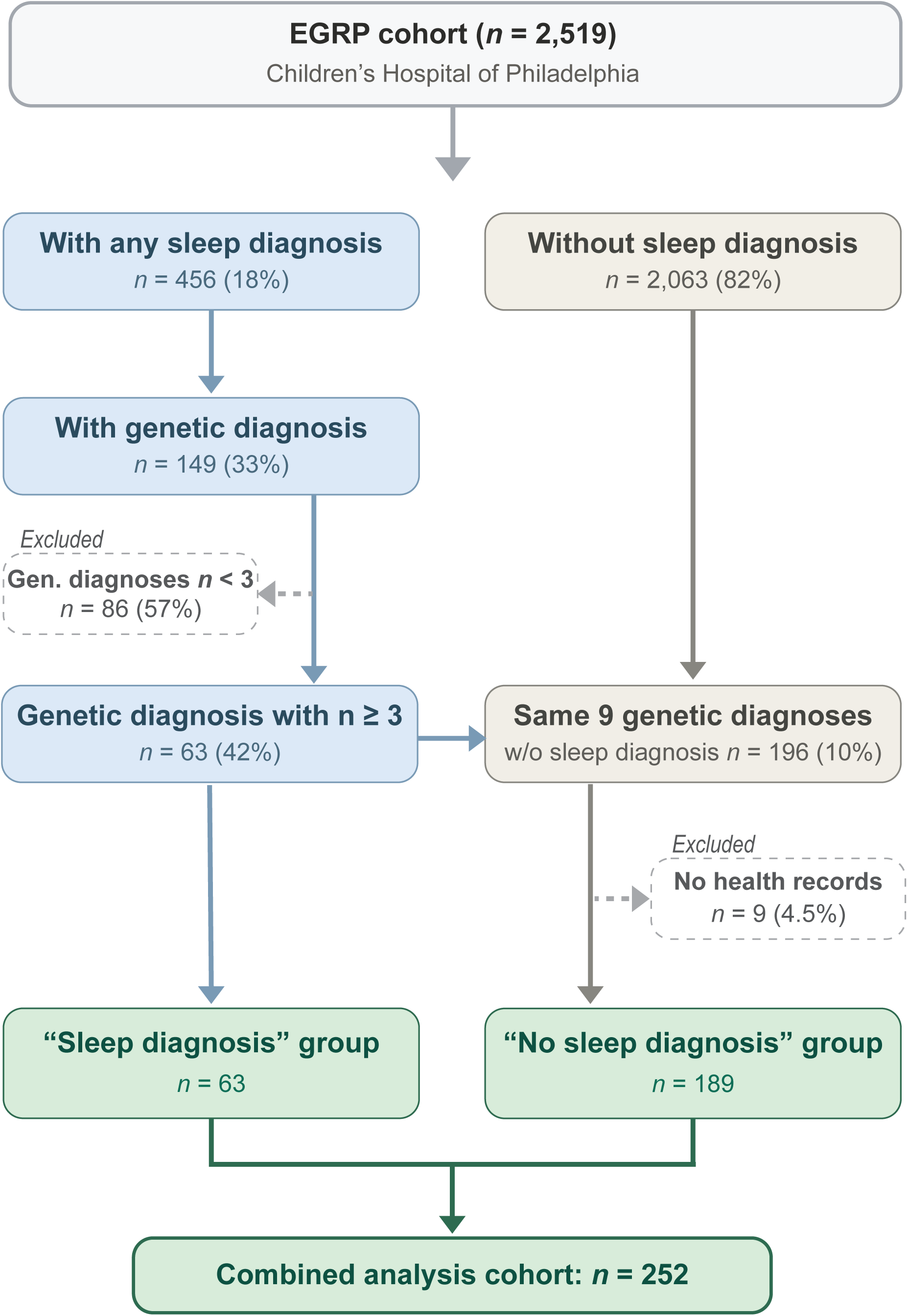
Derivation of the study cohort. Flowchart depicting stepwise selection of the analysis cohort from the Epilepsy Genetics Research Project (EGRP) at the Children’s Hospital of Philadelphia. Of 2,519 individuals with neurodevelopmental disorders, 456 (18%) carried a formal sleep-related diagnosis, of whom 149 had a confirmed genetic aetiology. The nine genetic diagnoses most frequently associated with a sleep-related diagnosis in this cohort (each occurring in three or more individuals) formed the sleep diagnosis group (*n* = 63). A subsequent analysis identified individuals with the same nine genetic diagnoses but without a sleep-related diagnosis (no sleep diagnosis group; *n* = 189), yielding a combined analysis cohort of 252 individuals.

**Table 1.** Demographics of study cohort including genetic diagnosis, sleep disorders, and neuropsychiatric comorbidities.

| Group |  | Demographics |  |  | Neuropsychiatric comorbidities |  |  |
| --- | --- | --- | --- | --- | --- | --- | --- |
| Gene | Sleep disorder diagnosis | N (%) | Average age (years) | Female n/N (%) | ASD n/N (%) | ADHD n/N (%) | Mood disorder n/N (%) |
| SCN1A | Presence | 22 (39) | 9 | 11/22 (50%) | 4/22 (18%) | 7/22 (32%) | 2/22 (9%) |
|  | Absence | 35 (61) | 9.1 | 19/35 (54%) | 8/35 (23%) | 2/35 (6%) | 3/35 (9%) |
| SCN2A | Presence | 7 (21) | 7.1 | 3/7 (43%) | 7/7 (100%) | 1/7 (14%) | 1/7 (14%) |
|  | Absence | 27 (79) | 8.7 | 13/27 (48%) | 7/27 (26%) | 0/27 (0%) | 1/27 (4%) |
| STXBP1 | Presence | 11 (14) | 7.9 | 5/11 (45%) | 4/11 (36%) | 1/11 (9%) | 0/11 (0%) |
|  | Absence | 68 (86) | 9.7 | 32/68 (47%) | 10/68 (15%) | 3/68 (4%) | 4/68 (6%) |
| WDR45 | Presence | 4 (31) | 9.2 | 3/4 (75%) | 3/4 (75%) | 1/4 (25%) | 2/4 (50%) |
|  | Absence | 9 (69) | 13.9 | 8/9 (89%) | 6/9 (67%) | 1/9 (11%) | 1/9 (11%) |
| SLC6A1 | Presence | 3 (21) | 11.7 | 0/3 (0%) | 2/3 (67%) | 0/3 (0%) | 1/3 (33%) |
|  | Absence | 11 (79) | 12.1 | 5/11 (45%) | 1/11 (9%) | 1/11 (9%) | 0/11 (0%) |
| KCNQ2 | Presence | 4 (19) | 5.8 | 1/4 (25%) | 0/4 (0%) | 0/4 (0%) | 0/4 (0%) |
|  | Absence | 17 (81) | 6.4 | 7/17 (41%) | 1/17 (6%) | 0/17 (0%) | 1/17 (6%) |
| KCNT1 | Presence | 5 (45) | 8.2 | 3/5 (60%) | 2/5 (40%) | 1/5 (20%) | 0/5 (0%) |
|  | Absence | 6 (55) | 14.8 | 2/6 (33%) | 0/6 (0%) | 0/6 (0%) | 0/6 (0%) |
| PCDH19 | Presence | 4 (40) | 11.5 | 4/4 (100%) | 3/4 (75%) | 1/4 (25%) | 0/4 (0%) |
|  | Absence | 6 (60) | 9.3 | 6/6 (100%) | 3/6 (50%) | 0/6 (0%) | 3/6 (50%) |
| SYNGAP1 | Presence | 3 (23) | 10.7 | 1/3 (33%) | 2/3 (67%) | 1/3 (33%) | 1/3 (33%) |
|  | Absence | 10 (77) | 8.9 | 5/10 (50%) | 6/10 (60%) | 1/10 (10%) | 1/10 (10%) |

Abbreviations. ASD, autism spectrum disorder; ADHD, attention deficit hyperactivity disorder.
“Mood disorder” includes depression and/or anxiety-related disorders.
N is the total number of individuals associated within a specific category and type of finding. In the remaining columns, values are presented as n/N (%), which represents the number of cases (n) that fall under the given category out of the total number (N), expressed as a percentage. All percentages are rounded to the nearest percent. Average ages are rounded to the nearest tenth.

**Table 2.**
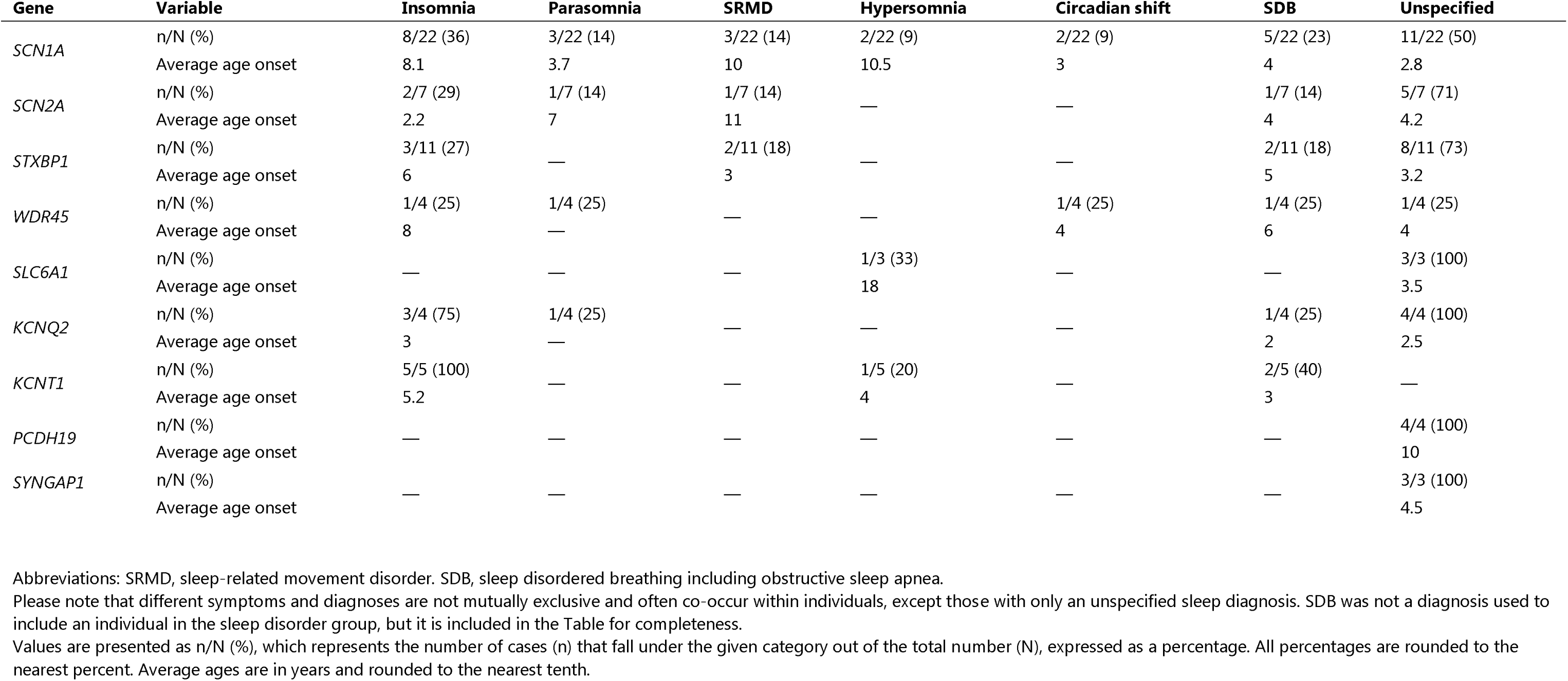
Characteristics of sleep disorder diagnoses for monogenic etiologies of epilepsy.

| Gene | Variable | Insomnia | Parasomnia | SRMD | Hypersomnia | Circadian shift | SDB | Unspecified |
| --- | --- | --- | --- | --- | --- | --- | --- | --- |
| *SCN1A* | n/N (%) | 8/22 (36) | 3/22 (14) | 3/22 (14) | 2/22 (9) | 2/22 (9) | 5/22 (23) | 11/22 (50) |
|  | Average age onset | 8.1 | 3.7 | 10 | 10.5 | 3 | 4 | 2.8 |
| *SCN2A* | n/N (%) | 2/7 (29) | 1/7 (14) | 1/7 (14) | — | — | 1/7 (14) | 5/7 (71) |
|  | Average age onset | 2.2 | 7 | 11 | — | — | 4 | 4.2 |
| *STXBP1* | n/N (%) | 3/11 (27) | — | 2/11 (18) | — | — | 2/11 (18) | 8/11 (73) |
|  | Average age onset | 6 | — | 3 | — | — | 5 | 3.2 |
| *WDR45* | n/N (%) | 1/4 (25) | 1/4 (25) | — | — | 1/4 (25) | 1/4 (25) | 1/4 (25) |
|  | Average age onset | 8 | — | — | — | 4 | 6 | 4 |
| *SLC6A1* | n/N (%) | — | — | — | 1/3 (33) | — | — | 3/3 (100) |
|  | Average age onset | — | — | — | 18 | — | — | 3.5 |
| *KCNQ2* | n/N (%) | 3/4 (75) | 1/4 (25) | — | — | — | 1/4 (25) | 4/4 (100) |
|  | Average age onset | 3 | — | — | — | — | 2 | 2.5 |
| *KCNT1* | n/N (%) | 5/5 (100) | — | — | 1/5 (20) | — | 2/5 (40) | — |
|  | Average age onset | 5.2 | — | — | 4 | — | 3 | — |
| *PCDH19* | n/N (%) | — | — | — | — | — | — | 4/4 (100) |
|  | Average age onset | — | — | — | — | — | — | 10 |
| *SYNGAP1* | n/N (%) | — | — | — | — | — | — | 3/3 (100) |
|  | Average age onset | — | — | — | — | — | — | 4.5 |

Abbreviations: SRMD, sleep-related movement disorder. SDB, sleep disordered breathing including obstructive sleep apnea.
Please note that different symptoms and diagnoses are not mutually exclusive and often co-occur within individuals, except those with only an unspecified sleep diagnosis. SDB was not a diagnosis used to include an individual in the sleep disorder group, but it is included in the Table for completeness.
Values are presented as n/N (%), which represents the number of cases (n) that fall under the given category out of the total number (N), expressed as a percentage. All percentages are rounded to the nearest percent. Average ages are in years and rounded to the nearest tenth.

### Manual data retrieval from electronic medical record (EMR)

Clinical information was manually extracted by two reviewers (A.I.B. and V.A.C.). Demographic and background information were obtained from a combination of keyword searches in the EMR and the main problem list. Subjective sleep-related data including self-reported sleep symptoms, bedtimes, sleep medications and sleep response to medications, as well as epilepsy-related data including sleep/night associated seizures were obtained from targeted keyword searches. Polysomnography (PSG) data were available for a limited number of individuals, and it was thus excluded from the overall analysis. We note that individuals’ PSG records often confirmed subjective sleep findings.

Seizure management was assessed using a bespoke six-tier grading system, ranging from no lifetime seizures (Grade 0) to poorly controlled epilepsy (Grade 5). A Grade of 1 or 2 was given to patients who did not have a seizure in the year before their most recent neurology visit based on whether they were currently off (Grade 1) or on (Grade 2) an anti-seizure medication (ASM). Those on ASMs with ongoing seizures were given a Grade 3 if seizures were controlled at a stable/consistent rate, a Grade 4 was given if the seizure rate was mixed or unclear and a Grade 5 was given if the rate worsened at the most recent neurology visit. We then used these grades to develop the following seizure outcome categories: Seizure free = Grades 0 to 2, stable rate of seizures = Grade 3 and poor seizure control = Grades 4 and 5. Sleep medication success was determined by whether qualitative improvement in symptoms was documented in notes following the implementation of the regimen.

### Data analysis

To describe sleep phenotypes associated with monogenic epilepsies, we identified (1) the type and frequency of sleep diagnoses (retrieved algorithmically), (2) self-reported sleep symptoms (retrieved manually) and (3) EEG characterization of sleep architecture (retrieved manually), taking note of the description of disrupted, abnormal, or lack of sleep architecture and presence of abnormal waveforms during sleep. Many individuals with formal sleep diagnoses also had documented sleep symptoms, and conversely, many without formal diagnoses had documented symptoms Combining the “sleep diagnosis” and “no sleep diagnosis” groups enabled us to understand the sleep-related symptoms and endophenotypes associated with monogenic diagnoses, while comparing these two groups for each genetic diagnosis allowed us to characterize the extent of underdiagnosis of sleep disorders.

A table of data retrieved from the EMR was used to derive the descriptive statistics found in Tables 1 to 5 and all inferential models. Among those with a sleep-related diagnosis, there was one individual with pathogenic variants in both *KCNT1* and *PCDH19* genes. When data were analyzed as a function of genetic diagnosis, the former individual’s data were counted twice, once for each gene. Among those without a sleep-related diagnosis, there was one individual with pathogenic variants in both *SCN2A* and *STXBP1* whose data were treated in the same manner. Seven individuals in the "no sleep diagnosis" group (one each for *KCNQ2*, *KCNT1* and *PCDH19* and four for *STXBP1*) whose EMR did not contain clinical information beyond their monogenic diagnosis were excluded from the study entirely. Denominators vary across analyses and reflect the number of individuals with available data for each outcome, consistent with the variable completeness of electronic medical record documentation. Gene-stratified results are presented as counts and proportions and are not accompanied by formal inferential tests, given small within-gene subgroup sizes. For *KCNQ2*-related disorders, individuals were further classified by clinical subtype (self-limited familial neonatal epilepsy or neonatal-onset developmental and epileptic encephalopathy) based on documented clinical diagnosis in the EMR; three individuals whose records did not permit subtype classification were excluded from this specific analysis.

Seizure persistence was defined from the six-tier seizure management severity grade described in the section above as Grade ≥ 3 (persistent) versus Grade < 3 (nonpersistent).

Inferential analyses used Bayesian logistic regression with weakly informative priors (Normal(0, 2.5) for the intercept; Normal(0, 1) for predictor coefficients), rather than conventional frequentist tests, to avoid the estimation instability that arises from sparse or zero cells in uncommon sleep subtypes. Results are reported as posterior median odds ratios (*OR*s) with 95% credible intervals (*CrI*s); an association was considered credible when the *95% CrI* excluded *OR* = 1.

To examine whether persistent seizures were associated with greater sleep-related phenotype burden, separate Bayesian logistic regression models were fitted with persistent seizure status as the binary predictor and each sleep-related phenotype as a binary outcome, first examining domain-level composite outcomes (Figure 3A) and then individual sleep subtypes and EEG features in exploratory post-hoc models (Figure 3B). To examine whether neuropsychiatric comorbidities independently predicted sleep burden, the same modelling approach was applied with autism spectrum disorder (ASD), attention-deficit/hyperactivity disorder (ADHD), and mood disorders (anxiety and depression) as the predictor and either sleep disorder diagnosis or any sleep issue as the outcome (Figure 3D); models were repeated with adjustment for persistent seizure status to assess independence from seizure severity. Gene-stratified prevalence differences are presented descriptively in Figure 3C. All models were fitted using the brms package in R version 4.5.2.

## Results

### Prevalence of under-recognition of sleep dysfunction

Of 2,519 individuals with neurodevelopmental disorders enrolled in the EGRP, 456 (18%) carried a sleep-related diagnosis (Figure 1). Of these, 149 (33%) had a confirmed genetic aetiology. Individuals with one of the nine genetic diagnoses most frequently associated with a sleep-related diagnosis (each occurring three or more times) formed the “sleep diagnosis” group (*n=*63). Parallel analysis identified individuals with the same nine genetic diagnoses but without a sleep-related diagnosis (*n=*189), yielding a combined analysis cohort of 252 individuals. We identified these nine genes as most frequently associated with both epilepsy and a sleep-related diagnosis: *SCN1A*, *STXBP1*, *SCN2A*, *KCNT1*, *PCDH19*, *KCNQ2*, *SLC6A1*, *SYNGAP1* and *WDR45* (Table 1). Gene-level cohort sizes ranged from 10 (*PCDH19*) to 79 (*STXBP1*). The genetic aetiologies with the highest proportion of individuals carrying a formal sleep diagnosis were *KCNT1* (45%, 5/11), *PCDH19* (40%, 4/10) and *SCN1A* (39%, 22/57), although sample sizes varied considerably across genes.

Formal sleep diagnoses captured only a fraction of the true sleep burden in this cohort (Figure 2A). Among the 189 individuals without a formal sleep diagnosis, 82 (43%) had sleep-related symptoms documented in the medical record, most commonly insomnia (21%, *n=*39), sleep-disordered breathing features (19%, *n=*36) and hypersomnolence (12%, *n=*22). When symptom reports were included alongside formal diagnoses, 145 of 252 individuals (58%) had documented sleep involvement, more than double the rate captured by diagnostic coding alone (25%, 63/252). This pattern was evident across all nine genetic diagnoses, with the highest rates of symptom burden among undiagnosed individuals observed in *SLC6A1* (64%, 7/11), *WDR45* (56%, 5/9) and *SCN1A* (51%, 18/35; Figure 2A). The true sleep burden may be higher still: Symptom documentation rates were consistently elevated among individuals with formal diagnoses compared to those without across all domains (e.g. insomnia symptoms: 79% vs 21%; Table 3), suggesting that clinical documentation practices may differ systematically between these groups, with sleep complaints potentially underreported in individuals lacking a formal sleep diagnosis.

**Figure 2.**
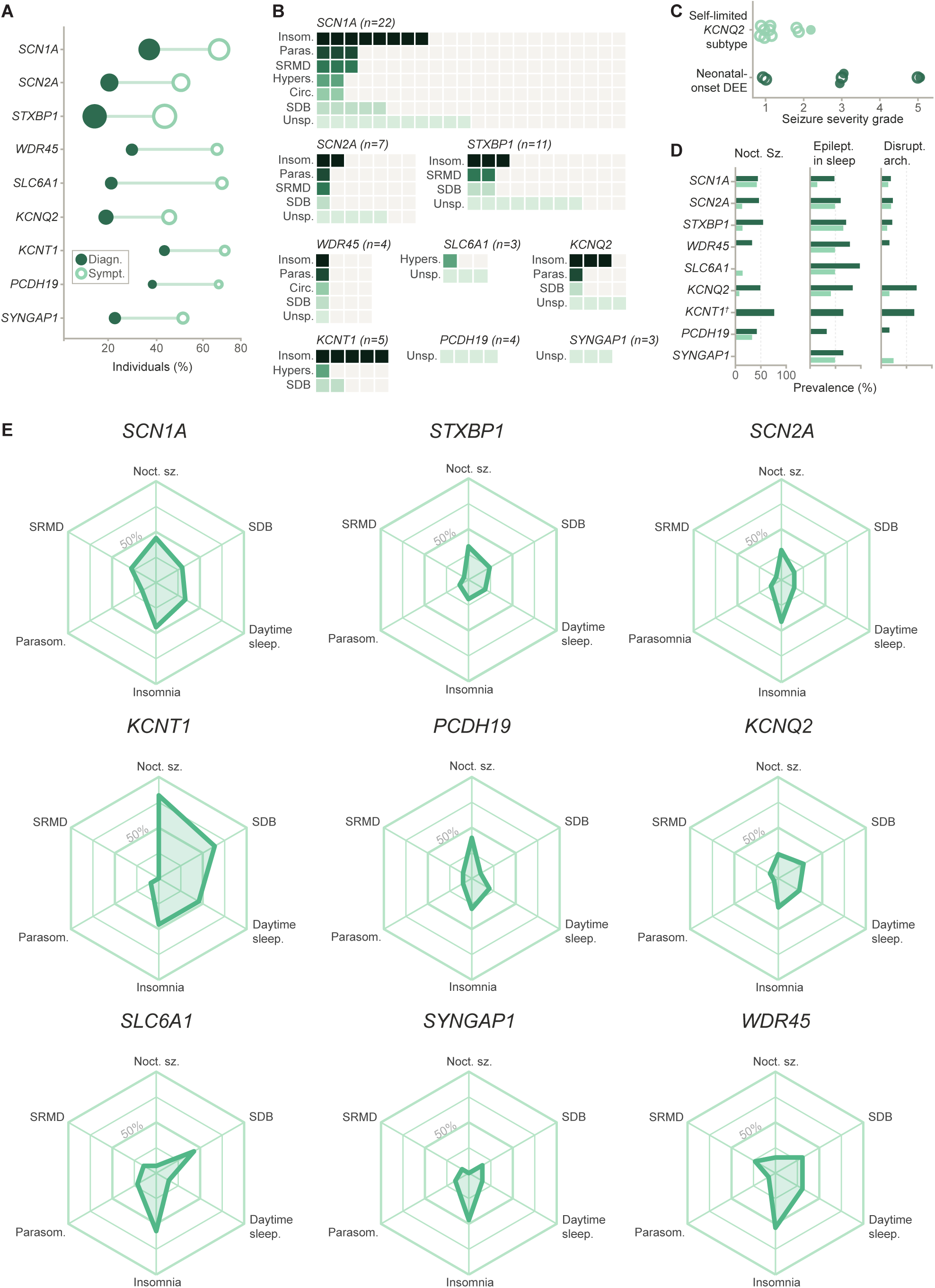
Sleep phenotype prevalence and phenotype profiles across monogenic epilepsies. **(A)** Prevalence of formal sleep diagnosis and any sleep involvement per genetic diagnosis. Filled circles indicate the proportion of individuals with a formal sleep disorder diagnosis; open circles indicate the proportion with any documented sleep involvement, defined as a formal diagnosis or a self-reported sleep symptom abstracted from clinical notes. Circle size is proportional to the total number of individuals per genetic diagnosis. **(B)** Formal sleep diagnosis profiles within the sleep diagnosis group, shown separately for each genetic diagnosis. Each column represents one individual with a formal sleep diagnosis; each row represents one diagnosis type. Filled squares indicate the number of individuals within that gene group who received that diagnosis, sorted left to right within each row; empty outlined squares indicate individuals without that diagnosis. Diagnosis types absent for a gene indicate no individual in that group received that diagnosis. Colour encodes diagnosis type from darkest to lightest: insomnia, parasomnia, sleep-related movement disorder, hypersomnolence, circadian dysfunction, sleep-disordered breathing and unspecified. A single individual may have more than one diagnosis and filled squares across different rows do not imply the same individual. **(C)** Seizure severity grade for individuals with KCNQ2-related disorders, stratified by subtype. Each circle represents one individual. Dark green indicates neonatal-onset developmental and epileptic encephalopathy; light green indicates the self-limited subtype. Filled circles indicate a formal sleep diagnosis; open circles indicate no formal sleep diagnosis. **(D)** Prevalence of nocturnal seizures, epileptiform activity during sleep and disrupted sleep architecture on EEG across genetic diagnoses, stratified by seizure persistence (light green: nonpersistent, Grades 0–2; dark green: persistent, Grades 3–5). Each panel shows the percentage of individuals per gene with that feature. Denominators for nocturnal seizures reflect all individuals with available data per gene; denominators for epileptiform activity and disrupted sleep architecture reflect only individuals with EEG documentation capturing sleep. †*KCNT1*: all seizure-graded individuals had persistent seizures (*n* = 9; 2 ungradable), precluding a nonpersistent comparison for this gene. **(E)** Radar plots display the prevalence of six sleep-related features for each of the nine genetic diagnoses across the full analysis cohort (*n* = 252). Each axis is scaled from 0 to 100%, with the 50% reference ring labelled. Six features are shown clockwise from top: nocturnal or sleep–wake transition seizures (Noct. sz.), sleep-disordered breathing (SDB), daytime sleepiness (Daytime sleepin.), insomnia, parasomnia (Parasom.) and sleep-related movement disorder (SRMD). Phenotypes are not mutually exclusive.

**Table 3.** Self-reported sleep symptoms and sleep EEG in individuals across monogenic diagnoses and sleep groups.

| Gene | Sleep disorder diagnosis | Insomnia | Nightmares or terrors | Other parasomnias | SRMD | SDB | Daytime sleepiness | Noct./sleep-wake seizures | Disrupted sleep arch<sup>a</sup> | Epileptiform in sleep<sup>a</sup> |
| --- | --- | --- | --- | --- | --- | --- | --- | --- | --- | --- |
| SCN1A | Presence | 17/22 (77) | 1/22 (5) | 4/22 (18) | 10/22 (45) | 10/22 (45) | 16/22 (73) | 8/22 (36) | 7/21 (33) | 14/20 (70) |
|  | Absence | 8/35 (23) | 1/35 (3) | 3/35 (9) | 6/35 (17) | 7/35 (20) | 3/35 (9) | 17/35 (49) | 4/33 (12) | 9/31 (29) |
|  | Overall | 25/57 (44) | 2/57 (4) | 7/57 (12) | 16/57 (28) | 17/57 (30) | 19/57 (33) | 25/57 (44) | 11/54 (20) | 23/51 (45) |
| SCN2A | Presence | 7/7 (100) | 1/7 (14) | 1/7 (14) | 2/7 (29) | 2/7 (29) | 2/7 (29) | 3/7 (43) | 1/7 (14) | 5/7 (71) |
|  | Absence | 7/27 (26) | 1/27 (4) | 1/27 (4) | 0/27 (0) | 3/27 (11) | 3/27 (11) | 7/27 (26) | 7/27 (26) | 9/18 (50) |
|  | Overall | 14/34 (41) | 2/34 (6) | 2/34 (6) | 2/34 (6) | 5/34 (15) | 5/34 (15) | 10/34 (29) | 8/34 (24) | 14/25 (56) |
| STXBP1 | Presence | 6/11 (55) | 0/11 (0) | 3/11 (27) | 3/11 (27) | 7/11 (64) | 7/11 (64) | 5/11 (45) | 3/10 (30) | 8/10 (80) |
|  | Absence | 9/68 (13) | 1/68 (1) | 5/68 (7) | 1/68 (1) | 12/68 (18) | 8/68 (12) | 21/68 (31) | 10/67 (15) | 29/43 (67) |
|  | Overall | 15/79 (19) | 1/79 (1) | 8/79 (10) | 4/79 (5) | 19/79 (24) | 15/79 (19) | 26/79 (33) | 13/77 (17) | 37/53 (70) |
| WDR45 | Presence | 4/4 (100) | 1/4 (25) | 0/4 (0) | 2/4 (50) | 2/4 (50) | 2/4 (50) | 1/4 (25) | 0/3 (0) | 3/3 (100) |
|  | Absence | 3/9 (33) | 0/9 (0) | 0/9 (0) | 1/9 (11) | 2/9 (22) | 2/9 (22) | 1/9 (11) | 1/9 (11) | 2/4 (50) |
|  | Overall | 7/13 (54) | 1/13 (8) | 0/13 (0) | 3/13 (23) | 4/13 (31) | 4/13 (31) | 2/13 (15) | 1/12 (8) | 5/7 (71) |
| SLC6A1 | Presence | 3/3 (100) | 0/3 (0) | 2/3 (67) | 1/3 (33) | 3/3 (100) | 1/3 (33) | 0/3 (0) | 0/3 (0) | 3/3 (100) |
|  | Absence | 5/11 (45) | 1/11 (9) | 1/11 (9) | 1/11 (9) | 3/11 (27) | 1/11 (9) | 1/11 (9) | 0/11 (0) | 4/9 (44) |
|  | Overall | 8/14 (57) | 1/14 (7) | 3/14 (21) | 2/14 (14) | 6/14 (43) | 2/14 (14) | 1/14 (7) | 0/14 (0) | 7/12 (58) |
| KCNQ2 | Presence | 3/4 (75) | 1/4 (25) | 0/4 (0) | 2/4 (50) | 2/4 (50) | 3/4 (75) | 3/4 (75) | 3/4 (75) | 4/4 (100) |
|  | Absence | 3/17 (18) | 0/17 (0) | 0/17 (0) | 0/17 (0) | 4/17 (24) | 2/17 (12) | 2/17 (12) | 4/15 (27) | 7/15 (47) |
|  | Overall | 6/21 (29) | 1/21 (5) | 0/21 (0) | 2/21 (10) | 6/21 (29) | 5/21 (24) | 5/21 (24) | 7/19 (37) | 11/19 (58) |
| KCNT1 | Presence | 5/5 (100) | 0/5 (0) | 1/5 (20) | 0/5 (0) | 4/5 (80) | 3/5 (60) | 4/5 (80) | 4/5 (80) | 3/4 (75) |
|  | Absence | 0/6 (0) | 0/6 (0) | 0/6 (0) | 0/6 (0) | 3/6 (50) | 2/6 (33) | 5/6 (83) | 2/6 (33) | 1/2 (50) |
|  | Overall | 5/11 (45) | 0/11 (0) | 1/11 (9) | 0/11 (0) | 7/11 (64) | 5/11 (45) | 9/11 (82) | 6/11 (55) | 4/6 (67) |
| PCDH19 | Presence | 2/4 (50) | 0/4 (0) | 1/4 (25) | 0/4 (0) | 0/4 (0) | 1/4 (25) | 1/4 (25) | 1/3 (33) | 2/4 (50) |
|  | Absence | 1/6 (17) | 0/6 (0) | 0/6 (0) | 1/6 (17) | 1/6 (17) | 1/6 (17) | 3/6 (50) | 0/6 (0) | 3/4 (75) |
|  | Overall | 3/10 (30) | 0/10 (0) | 1/10 (10) | 1/10 (10) | 1/10 (10) | 2/10 (20) | 4/10 (40) | 1/9 (11) | 5/8 (62) |
| SYNGAP1 | Presence | 3/3 (100) | 0/3 (0) | 1/3 (33) | 1/3 (33) | 1/3 (33) | 0/3 (0) | 0/3 (0) | 0/2 (0) | 0/3 (0) |
|  | Absence | 3/10 (30) | 1/10 (10) | 0/10 (0) | 0/10 (0) | 1/10 (10) | 2/10 (20) | 0/10 (0) | 1/10 (10) | 4/7 (57) |
|  | Overall | 6/13 (46) | 1/13 (8) | 1/13 (8) | 1/13 (8) | 2/13 (15) | 2/13 (15) | 0/13 (0) | 1/12 (8) | 4/10 (40) |

Abbreviations. Arch, architecture; SRMD, sleep-related movement disorder;; SDB, sleep disordered breathing includes apnea diagnosis.
Please note that different symptoms and diagnoses are not mutually exclusive and often co-occur within individuals.
Values are presented as n/N (%), which represents the number of cases (n) that fall under the given category out of the total number (N), expressed as a percentage. All percentages are rounded to the nearest percent.
<sup>a</sup>Measured by EEG

Pharmacological sleep management was more common, as expected, among individuals with a formal sleep diagnosis. Melatonin was prescribed to 33% of the cohort overall (83/252), with clonidine (10%, 25/252) and trazodone (2%, 5/252) used less frequently (Table 4). Sleep medications were used by 75% (47/63) of individuals with a formal sleep diagnosis versus 26% (48/189) of those without (*OR* 7.78, *95% CrI* 4.21–14.46), driven primarily by melatonin (70% vs 21%; *OR* 7.38, *95% CrI* 4.06–13.63) and clonidine (21% vs 6%; *OR* 3.10, *95% CrI* 1.41–6.75). Documentation of medication response was sparse and non-standardized, precluding systematic efficacy analysis.

**Table 4.** Medications improving sleep across monogenic diagnoses.

| Gene | Sleep diagnosis | Any sleep medication use, n/N (%) | Melatonin |  | Clonidine |  | Trazodone |  |
| --- | --- | --- | --- | --- | --- | --- | --- | --- |
|  |  |  | Use (%) | Improved (%) | Use (%) | Improved (%) | Use (%) | Improved (%) |
| *SCN1A* | Presence | 15/22 (68) | 15/15 (100) | 1/6 (17) | 3/15 (20) | 0/1 (0) | 2/15 (13) | 0/2 (0) |
|  | Absence | 11/35 (31) | 9/11 (82) | 0/1 (0) | 2/11 (18) | — | — | — |
| *SCN2A* | Presence | 5/7 (71) | 4/5 (80) | 2/2 (100) | 1/5 (20) | — | — | — |
|  | Absence | 10/27 (37) | 9/10 (90) | — | 3/10 (30) | 2/2 (100) | — | — |
| *STXBP1* | Presence | 5/11 (45) | 2/5 (40) | — | 3/5 (60) | 0/1 (0) | 1/5 (20) | 1/1 (100) |
|  | Absence | 13/68 (19) | 12/13 (92) | 1/2 (50) | 2/13 (15) | 1/1 (100) | 1/13 (8) | — |
| *WDR45* | Presence | 4/4 (100) | 4/4 (100) | 1/2 (50) | 2/4 (50) | 0/1 (0) | 1/4 (25) | 0/1 (0) |
|  | Absence | 4/9 (44) | 4/4 (100) | 1/1 (100) | — | — | — | — |
| *SLC6A1* | Presence | 3/3 (100) | 3/3 (100) | 1/1 (100) | — | — | — | — |
|  | Absence | 3/11 (27) | 1/3 (33) | — | 3/3 (100) | 1/1 (100) | — | — |
| *KCNQ2* | Presence | 4/4 (100) | 4/4 (100) | 2/4 (50) | 1/4 (25) | 1/1 (100) | — | — |
|  | Absence | 1/17 (6) | 1/1 (100) | — | — | — | — | — |
| *KCNT1* | Presence | 5/5 (100) | 5/5 (100) | 1/1 (100) | 2/5 (40) | 0/1 (0) | — | — |
|  | Absence | 0/6 (0) | — | — | — | — | — | — |
| *PCDH19* | Presence | 4/4 (100) | 4/4 (100) | 1/1 (100) | — | — | — | — |
|  | Absence | 2/6 (33) | 2/2 (100) | — | — | — | — | — |
| *SYNGAP1* | Presence | 3/3 (100) | 3/3 (100) | 2/2 (100) | 1/3 (33) | 1/1 (100) | — | — |
|  | Absence | 3/10 (30) | 1/3 (33) | — | 2/3 (67) | 1/2 (50) | — | — |

Note: different symptoms and diagnoses are not mutually exclusive and often co-occur within individuals.
Values are presented as n/N (%), which represents the number of cases (n) that fall under the given category out of the total number (N), expressed as a percentage. All percentages are rounded to the nearest percent.

### Sleep phenotype profiles vary by genetic aetiology

Each of the nine monogenic epilepsies in our cohort showed a distinctive sleep phenotype profile, characterized by gene-specific patterns of formal diagnoses, symptom burden, nocturnal seizures and EEG sleep features (Figure 2B-E).

*SCN1A*-related diagnoses showed the broadest spectrum of sleep phenotypes in this cohort. A formal sleep diagnosis was recorded in 39% of individuals (22/57), while sleep-related symptoms or diagnoses were documented in 70% (40/57). Among the 22 individuals with a formal sleep diagnosis (Figure 2B), insomnia was the most common specific diagnosis (36%, 8/22), alongside parasomnia (14%, 3/22) and sleep-related movement disorder (14%, 3/22); half carried an unspecified sleep disorder diagnosis (50%, 11/22). Across the full *SCN1A* group (Figure 2E), insomnia symptoms and nocturnal or sleep-wake seizures were documented at identical rates (44%, 25/57 each), raising the possibility that the two are associated; daytime sleepiness, reported in 33% (19/57), may further reflect this relationship.

Sleep-related movement disorder symptoms (28%, 16/57) were also prominent. Among individuals with EEG documentation, disrupted sleep architecture was observed in 20% (11/54) and epileptiform activity during sleep in 45% (23/51; Figure 2D). The sleep endophenotype of *SCN1A* is therefore characterized by a co-occurrence of insomnia symptoms and nocturnal or sleep-wake seizures as its most distinctive feature.

*SCN2A*-related epilepsy was distinguished by the high prevalence of insomnia symptoms (41%, 14/34), which substantially exceeded the formal insomnia diagnosis rate of 6% (2/34). A formal sleep diagnosis was recorded in 21% of individuals (7/34), while any sleep involvement was documented in 53% (18/34). Within the formally diagnosed group (*n=*7), insomnia was recorded in 29% (2/7), parasomnia in 14% (1/7) and sleep-related movement disorder in 14% (1/7); the majority carried a non-specific sleep disorder diagnosis (71%, 5/7). Nocturnal or sleep-wake seizures were reported in 29% (10/34). Among individuals with EEG documentation, disrupted sleep architecture was observed in 24% (8/34) and epileptiform activity during sleep in a notable 56% (14/25). This endophenotype is thus characterized by a prominent insomnia symptom burden that is largely not captured by formal diagnostic coding, alongside a notable rate of epileptiform activity during sleep.

For *STXBP1*-related disorders, the largest group in this cohort (*n =* 79), formal sleep diagnoses were recorded in only 14% of individuals (11/79), while sleep-related symptoms were documented in 46% (36/79). The most common feature across the full group was nocturnal or sleep-wake seizures (33%, 26/79), followed by insomnia symptoms (19%, 15/79) and daytime sleepiness (19%, 15/79); sleep-related movement disorder symptoms were infrequent at the group level (5%, 4/79). Among the 11 individuals with any formal sleep diagnosis, insomnia symptoms were reported by over half (55%, 6/11), yet only 27% (3/11) had a formal insomnia diagnosis and the majority of diagnoses were non-specific (73%, 8/11). This cohort also had the second highest occurrence of epileptiform activity during sleep amongst all epilepsies analyzed at 70% (37/53), while disrupted sleep architecture was observed in 17% (13/77). Together, *STXBP1*-related disorders are characterized by insomnia with limited formal diagnostic capture and frequent epileptiform activity during sleep.

*WDR45*-related disorders showed a high overall burden of sleep involvement, with a formal sleep diagnosis recorded in 31% (4/13) and symptoms documented in 69% (9/13). Insomnia symptoms were the most prevalent feature across the full group (54%, 7/13) while only coded as a formal diagnosis in 8% (1/13). Nightmares or night terrors were not reported in any individual across the full group, distinguishing *WDR45-*related disorders from several others where parasomnia features were more common. Formal diagnosis rates against the full group were low: Insomnia was formally coded in 8% (1/13) compared to 54% (7/13) of patients reporting symptoms. Among individuals with EEG documentation, epileptiform activity during sleep was present in 71% (5/7), the highest rate amongst these monogenic epilepsies, while disrupted sleep architecture was observed in only 8% (1/12). This pattern positions insomnia symptoms and sleep-associated epileptiform activity as defining features of the *WDR45* endophenotype.

*SLC6A1*-related disorders showed the highest insomnia symptom prevalence of any gene in this cohort (57%, 8/14), yet no individual received a formal insomnia diagnosis. This parallels the broader underdiagnosis of sleep involvement in this group (21% formally diagnosed vs 71% with reported sleep symptoms). Nightmares or night terrors specifically were documented in 21% (3/14), one of the highest rates across these genes. Nocturnal or sleep-wake seizures were infrequent at the group level (7%, 1/14). Epileptiform activity during sleep was present in 58% (7/12) of the EEG-documented subset. The *SLC6A1* endophenotype is notable for a substantial insomnia symptom burden that was almost entirely uncaptured by formal diagnostic coding, alongside frequent epileptiform activity in sleep but no recorded sleep architecture disruption.

As expected, *KCNQ2*-related epilepsies showed sleep phenotypes that were strongly stratified by epilepsy subtypes. Among classifiable individuals, the self-limited familial neonatal epilepsy subtype comprised the majority and was associated with a relatively preserved sleep profile: All achieved seizure freedom, and formal sleep diagnosis rates, nocturnal seizure rates, and EEG sleep abnormalities were substantially lower in this subtype than in the neonatal-onset developmental and epileptic encephalopathy (DEE) subtype (Figure 2C). Across the full *KCNQ2* group, formal sleep diagnoses were recorded in 19% (4/21) while 48% (10/21) reported some form of sleep issue. Insomnia symptoms were reported in 29% of the full group (6/21) and daytime sleepiness in 24% (5/21). Among the four individuals with a formal sleep diagnosis, insomnia was the predominant diagnosis (75%, 3/4); all four also carried an unspecified sleep disorder diagnosis. Disrupted sleep architecture was observed in 37% (7/19) and epileptiform activity during sleep in 58% (11/19), both among the higher rates across genes in this cohort. Nocturnal or sleep-wake seizures were documented in 24% (5/21). The *KCNQ2* sleep endophenotype is largely attributable to the DEE subtype and characterized by insomnia symptoms and high rates of EEG sleep abnormalities, whereas individuals with the self-limited subtype exhibited few documented sleep symptoms or abnormalities.

*KCNT1*-related epilepsies exhibited the most severe sleep phenotype in this cohort. Formal sleep diagnoses were recorded in 45% of individuals (5/11), while sleep-related symptoms were reported by 73% (8/11). Insomnia symptoms and daytime sleepiness were each present in 45% of the group (5/11); notably, all individuals with a formal sleep diagnosis carried a formal insomnia diagnosis (100%, 5/5), underscoring the centrality of insomnia within this subgroup. Disrupted sleep architecture was observed in 55% of individuals with EEG data (6/11), the highest rate across all genes analyzed, and epileptiform activity during sleep was present in 67% of those with available EEGs (4/6). Parent-reported nocturnal or sleep–wake seizures were the most prevalent sleep-related feature (82%, 9/11). Collectively, the *KCNT1* endophenotype is defined by a pronounced convergence of nocturnal seizures, insomnia and disrupted sleep architecture, reflecting a profoundly dysregulated sleep–wake axis.

*PCDH19-*related epilepsy showed one of the higher rates of formal sleep diagnosis in this cohort (40%, 4/10), with reports of sleep features documented in 70% (7/10), yet the clinical characterization of sleep dysfunction remained largely non-specific. All four individuals with a formal sleep diagnosis carried exclusively a non-specific sleep disorder code. Disrupted sleep architecture was observed in 11% (1/9), while epileptiform activity during sleep was present in 62% (5/8).

*SYNGAP1*-related disorders showed a distinct phenotype defined by a complete absence of nocturnal or sleep-wake seizures, with a formal sleep diagnosis recorded in 23% (3/13) and reports of sleep features documented in 54% (7/13). All three individuals with a formal sleep diagnosis carried exclusively a non-specific sleep disorder code, although insomnia was the dominant symptom across the full group (46%, 6/13). Disrupted sleep architecture was observed in only one patient (1/12, 8%), while epileptiform activity during sleep was present in 40% (4/10).

### Persistent seizures are associated with greater sleep and nocturnal seizure burden

Across genetic diagnoses, persistent seizures were associated with greater sleep burden, including seizures occurring during sleep or sleep–wake transitions (Figure 3A). Sleep disorder diagnoses were documented more often in individuals with persistent seizures than in those without (34%, 49/143 vs 14%, 14/99), corresponding to higher odds of a recorded sleep disorder diagnosis (*OR* 2.87, *95% CrI* 1.57–5.36), as well as higher odds of any sleep issue, including reported symptoms (*OR* 2.18, *95% CrI* 1.32–3.66). Conversely, individuals with a sleep diagnosis had lower odds of seizure freedom across the full cohort (22%, 14/63 vs 47%, 85/179; *OR* 0.35, *95% CrI* 0.18–0.64). The strongest association was observed for nocturnal and sleep-wake seizures, which were nearly four and a half times more likely in individuals with persistent seizures (*OR* 4.47, *95% CrI* 2.50–8.28) and were reported in 46% of persistent cases (66/143) compared with 14% of nonpersistent cases (14/99). Among sleep subtypes (Figure 3B), persistent seizures were associated with insomnia (*OR* 1.80, *95% CrI* 1.06–3.05), hypersomnolence (*OR* 2.38, *95% CrI* 1.27–4.58) and disrupted sleep architecture on EEG (*OR* 2.06, *95% CrI* 1.08–4.16; Figure 3B). Evidence for an association with epileptiform activity during sleep was weaker at the cohort level (*OR* 1.52, *95% CrI* 0.85–2.77); in the subset with EEG documentation, epileptiform activity during sleep occurred in 61% of persistent cases (73/119) and 51% of nonpersistent cases (34/67). Sleep symptoms were associated with higher odds of disrupted sleep architecture (*OR* 2.06, *95% CrI* 1.08–4.16), and this relationship persisted when restricting to those without a formal sleep diagnosis (*OR* 3.01, *95% CrI* 1.34–7.15), indicating that objective sleep architecture disruption is more common in symptomatic individuals even when no formal diagnosis has been recorded.

**Figure 3.**
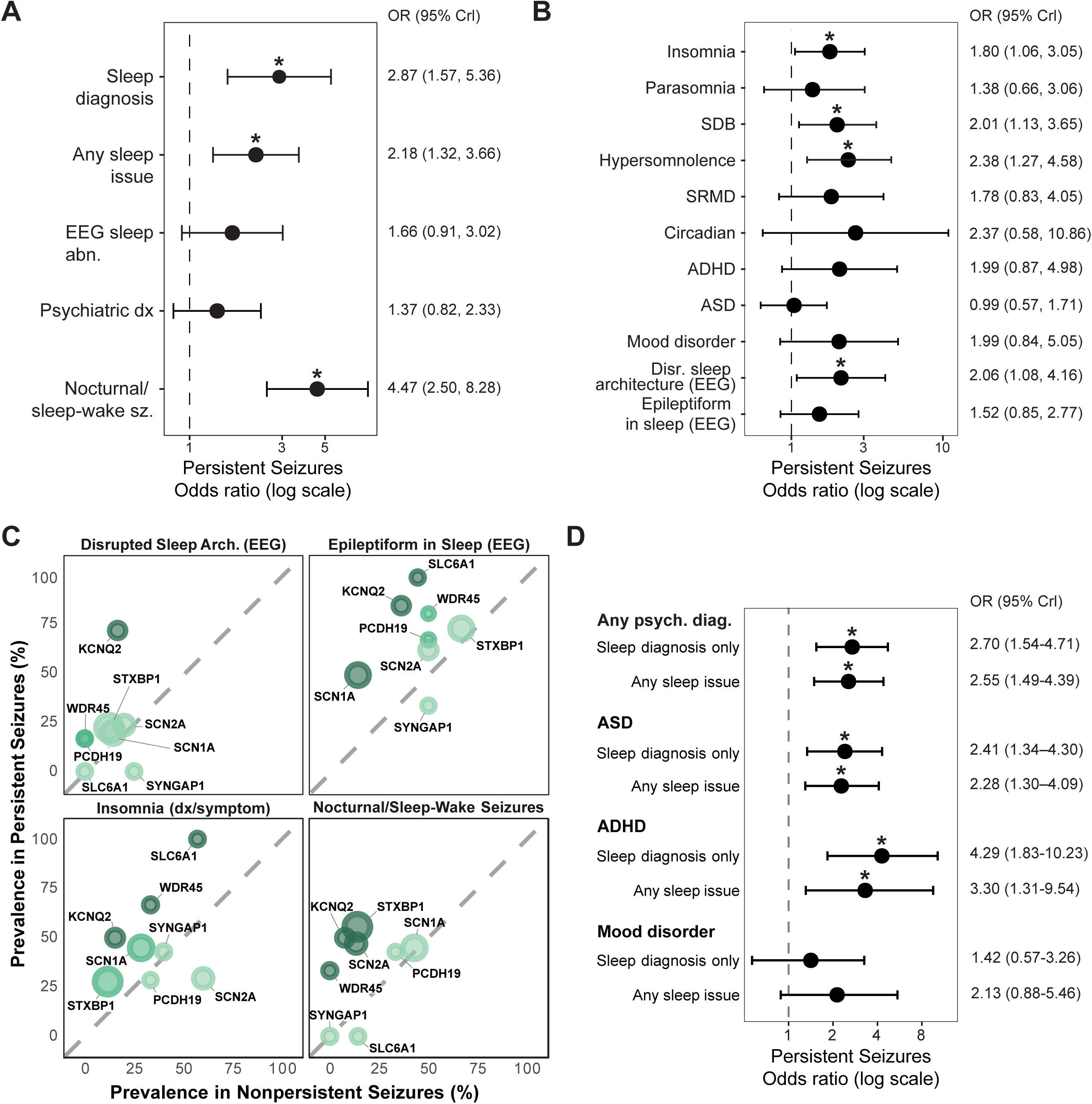
Associations between seizure persistence, neuropsychiatric comorbidities and sleep-related phenotypes across monogenic epilepsies. All models used Bayesian logistic regression with weakly informative priors; results are reported as posterior median odds ratios (*ORs*) with 95% credible intervals (*CrIs*). Asterisks denote associations where the *95% CrI* excludes OR = 1. **(A)** Domain-level associations with persistent seizures (Grade ≥ 3 vs. Grade < 3) as the predictor. Outcomes include formal sleep disorder diagnosis (Sleep dx), any sleep issue (diagnosis or documented symptom), any EEG sleep abnormality (comprising disrupted sleep architecture or epileptiform activity during sleep), any psychiatric diagnosis (ASD, ADHD, or mood disorder combined) and nocturnal or sleep–wake seizures. **(B)** Post-hoc breakdown of associations between persistent seizures and individual sleep subtypes and EEG features. Specific sleep subtypes reflect symptom-inclusive definitions (formal diagnosis or documented symptom report) unless otherwise indicated. Results should be interpreted with caution given smaller per-category sample sizes. **(C)** Gene-specific prevalence of four key sleep phenotypes by seizure persistence status. Each point represents one gene; x-axis shows prevalence in nonpersistent cases (Grade < 3) and y-axis shows prevalence in persistent cases (Grade ≥ 3). Points above the diagonal line (where *y = x*) indicate higher prevalence in the persistent group; distance from the diagonal reflects the magnitude of the severity-dependent gradient. Point size reflects total gene cohort size; colour indicates the magnitude of change (Δ% = persistent prevalence − nonpersistent prevalence). *KCNT1* excluded from scatter plots due to absence of individuals in the nonpersistent seizure group. **(D)** Associations between neuropsychiatric diagnoses and sleep burden, estimated separately for two sleep outcome definitions. For each neuropsychiatric predictor (any psychiatric diagnosis, ASD, ADHD and mood disorder), two models were run: one with formal sleep disorder diagnosis as the outcome (Sleep diagnosis only) and one with any sleep issue, defined as formal diagnosis or documented symptom report, as the outcome (Any sleep issue). Bold labels indicate the neuropsychiatric predictor group; indented labels indicate the sleep outcome definition. Mood disorder associations are shown for completeness; neither association reached credibility.

The strength of these associations varied substantially by genetic aetiology, revealing distinct endophenotypic patterns. Gene-level prevalence of nocturnal seizures, epileptiform activity during sleep, and disrupted sleep architecture across both seizure persistence groups is summarized in Figure 3C. *KCNQ2*-related disorders demonstrated the most consistent severity-dependent pattern, with substantially higher prevalence in persistent versus nonpersistent seizures across all four sleep outcomes: Disrupted sleep architecture (17% to 71%; the largest absolute difference observed), epileptiform activity during sleep (42% to 86%), insomnia (15% to 50%) and nocturnal seizures (8% to 50%). Other genes showed domain specific patterns. *STXBP1*- and *SCN2A*-related disorders exhibited strong associations between seizure persistence and nocturnal seizures (14% to 56% and 13% to 47%, respectively). *SYNGAP1*-related disorder showed consistently low rates of nocturnal seizures (0% in both groups) and disrupted sleep architecture regardless of seizure persistence, a pattern distinct from other aetiologies and consistent with its known role in sleep regulation^28^ rather than seizure-driven sleep disruption. *KCNQ2*-related disorders demonstrated the opposite pattern, with sleep phenotypes tightly tracking clinical severity: The self-limited familial neonatal epilepsy subtype (55% of classifiable cases, *n =* 10/18) showed uniformly preserved sleep profiles with 100% seizure freedom, minimal sleep diagnoses (10% vs 38% in neonatal-onset DEE), nocturnal seizures (10% vs 50%) and disrupted sleep architecture (0% vs 100%), while the neonatal-onset DEE subtype exhibited high burden across all sleep domains.

Taken together, these findings indicate that while persistent seizures are broadly associated with greater sleep burden across these monogenic epilepsies, the mechanisms linking sleep dysfunction to seizure severity vary substantially by genetic aetiology.

### Neuropsychiatric comorbidities contribute to sleep burden independent of seizure severity

Neuropsychiatric comorbidities were present in 88 of 252 individuals (35%) across the cohort, with ASD being the most prevalent (69/252, 27%), followed by ADHD and mood disorders (21/252, 8% each). Psychiatric diagnoses included in this study were associated with higher odds of a sleep disorder diagnosis (*OR* 2.70, *95% CrI* 1.54–4.71) and of documented sleep difficulties (*OR* 2.55, *95% CrI* 1.49–4.39; Figure 3D). Individually, ASD was associated with both sleep outcomes (sleep disorder diagnosis: *OR* 2.41, *95% CrI* 1.34–4.30; documented sleep difficulties: *OR* 2.28, *95% CrI* 1.30–4.09), as was ADHD (sleep disorder diagnosis: *OR* 4.29, *95% CrI* 1.83–10.23; documented sleep difficulties: *OR* 3.30, *95% CrI* 1.31–9.54), though credible intervals for ADHD were wide reflecting the small number of individuals with this diagnosis in the cohort (*n =* 21). Mood disorder did not show a credible association with either sleep diagnoses or reported symptoms. Importantly, associations between neuropsychiatric diagnoses and sleep burden persisted after adjustment for persistent seizure status (any psychiatric diagnosis: *OR* 2.49, *95% CrI* 1.40–4.49; ASD: *OR* 2.44, *95% CrI* 1.34–4.38; ADHD: *OR* 3.74, *95% CrI* 1.62–8.90), indicating that neuropsychiatric comorbidity and seizure severity contribute independently to sleep dysfunction rather than one simply reflecting the other.

At the gene level, notable patterns were observed (Table 1). *SCN2A*-related epilepsy showed the strongest association between ASD and sleep diagnosis, with all seven individuals carrying a formal sleep diagnosis also having ASD (100%) compared with 26% of those without a sleep diagnosis (7/27). *SLC6A1*-related disorders showed a similarly large difference for ASD (67%, 2/3 with sleep diagnosis vs 9%, 1/11 without). In *SCN1A*-related epilepsy, ADHD was more prevalent among individuals with a sleep disorder diagnosis (32%, 7/22) than those without (6%, 2/35), though all ADHD cases in this gene occurred in individuals with persistent seizures. *WDR45*-, *PCDH19*- and *SYNGAP1*-related disorders showed elevated neuropsychiatric comorbidity rates overall, with ASD present in 60–75% of individuals regardless of sleep diagnosis status.

## Discussion

Here we identify gene-specific sleep endophenotypes across nine monogenic epilepsies. These patterns suggest that sleep dysfunction in these disorders reflects distinct underlying pathophysiology rather than a uniform secondary consequence of seizure burden. Sleep difficulties were substantially underdiagnosed, and both seizure persistence and neuropsychiatric comorbidities contributed to overall sleep burden, indicating that multiple converging mechanisms drive sleep disruption in this population. These findings establish sleep as a possible therapeutic target for specific epilepsies.

We identified several previously under-unreported sleep phenotypes across monogenic epilepsies (Table 5). Parasomnia was observed in our cohort in *SCN1A*-, *SCN2A*-, *STXBP1*-, *WDR45*-, *SLC6A1*- *and KCNQ2*-related disorders, and sleep-related movement disorders in *SCN1A*-, *STXBP1*-, *PCDH19*- and *SYNGAP1*-related epilepsies. Hypersomnia symptoms were observed in *STXBP1*-, *KCNQ2*-, *PCDH19-* and *SYNGAP1*-related disorders. We also document disrupted sleep architecture on EEG in *STXBP1*-, *KCNQ2*-, *KCNT1*- *and SYNGAP1*-related disorders, sleep-related epileptiform activity in *SCN1A*-, *SCN2A*-, *SLC6A1*- and *KCNT1*-related epilepsies, and nocturnal seizures in *STXBP1*-related epilepsy. Circadian disturbance was identified in *WDR45*-related disorders. Prominent insomnia in *KCNT1*-related epilepsy may be associated with high rates of sleep-disordered breathing.

**Table 5.** Summary of sleep disorder diagnoses, neuropsychiatric comorbidities, patterns of epileptic activity in monogenic epilepsies.

| Condition | *SCN1A* | *SCN2A* | *STXBP1* | *WDR45* | *SLC6A1* | *KCNQ2* | *KCNT1* | *PCDH19* | *SYNGAP1* |
| --- | --- | --- | --- | --- | --- | --- | --- | --- | --- |
| Insomnia | ■ ■ ■<sup>30,31,50</sup> | ■ ■ ■<sup>32</sup> ◇<sup>51</sup> | ■ ■ | ■ ■ ■<sup>52</sup> | ■ | ■ ■ ■<sup>33</sup> ◇<sup>44</sup> | ■ | ■ ■<sup>15</sup> | ■ ■<sup>34,35</sup> |
| Parasomnia | ■ ■ | ■ ■ | ■ | ■ | ■ | ■ ■<sup>33</sup> | ■<sup>53</sup> |  | ■<sup>34,46</sup> ◇<sup>54</sup> |
| Sleep-related movement disorder | ■ ■ | ■ | ■ ■ | ■ | ■ |  |  | ■ ■ | ■ |
| Hypersomnia | ■ ■ ■<sup>30</sup> | ■ ■<sup>55</sup> | ■ | ■ ■<sup>52</sup> | ■ ■ ◇<sup>56,57</sup> | ■ | ■ ■ ◇<sup>58</sup> | ■ | ■ |
| Circadian disturbance | ■ ◇<sup>36,37</sup> | ◇<sup>39,40,51</sup> |  | ■ | ◇<sup>40</sup> |  |  | ◇<sup>41</sup> | ◇<sup>42</sup> |
| Unspecified sleep dysfunction | ■ | ■ | ■ ■<sup>59</sup> | ■ ■<sup>26</sup> | ■ ■<sup>29</sup> | ■ ■<sup>33</sup> |  | ■ ■<sup>15</sup> | ■ ◇<sup>60</sup> |
| Abnormal sleep architecture | ■ ■<sup>30,61</sup> ◇<sup>36,62,63</sup> | ■ ◇<sup>51</sup> | ■ | ■ ■<sup>52</sup> | ■<sup>64</sup> | ■ | ■ | ■ ■<sup>65</sup> | ■ |
| Sleep-related abnormal epileptic activity | ■ | ■ | ■ ■<sup>66</sup> | ■ ■<sup>67,68</sup> | ■ | ■ ■<sup>69</sup> | ■ | ■ ■<sup>70</sup> | ◇<sup>60,71</sup> |
| Nocturnal or sleep/wake-related seizures | ■ ■<sup>45</sup> ◇<sup>72,73</sup> | ■ ■<sup>73</sup> | ■ | ■ ■<sup>67</sup> | ■ ■<sup>74,75</sup> | ■ ■<sup>73,74</sup> | ■ ■<sup>76-78</sup> | ■ ■<sup>70</sup> | ■ ◇<sup>60</sup> |
| ASD | ■ | ■ ◇<sup>51</sup> | ■ ■<sup>79</sup> | ■ | ■ | ■ | ■ | ■ ■<sup>15</sup> | ■ ■<sup>35,47</sup> |
| ADHD | ■ ■<sup>80</sup> | ■ | ■ | ■ | ■ ■<sup>81</sup> |  | ■ | ■ | ■ ■<sup>47</sup> |
| Mood disorders | ■ ■<sup>80</sup> | ■ | ■ | ■ | ■ | ■ |  | ■ | ■ ■<sup>15,46</sup> |

■ = present in our study, ■ = documented symptom without formal diagnosis (our study), ■ = found in clinical literature, ◇ = found in preclinical literature

Our findings both validate and extend prior clinical reports. We confirm previously documented insomnia in *SCN1A*,^29–31^ *SCN2A*,^32^ WDR45,^35^ *KCNQ2*, ^33^ *PCDH19*,^15^ and *SYNGAP1*-related disorders,^34,35^ and circadian disturbance in *SCN1A*-related epilepsy.^36,37^ However, circadian dysfunction reported in the literature for *SCN2A*,^38,39^ *SLC6A1*,^40^ *PCDH19*,^41^ and *SYNGAP1*^42^ was not observed in our cohort. This likely reflects both the limitations of retrospective EMR-based phenotyping, which may under-ascertain more subtle symptoms, and our smaller sample sizes.

Disrupted sleep architecture on EEG was associated with both persistent seizures and documented sleep symptoms. This is significant because poor sleep has been associated with worse seizure control,^43^ and our findings indicate that objective sleep disruption can occur even in the absence of clinically recognized sleep disorders. Potassium channelopathies related to *KCNT1* and *KCNQ2* were especially likely to exhibit altered sleep architecture in our cohort. In mice, *Kcnq2*, which encodes for the voltage-gated potassium channel K_v_7.2, plays an important role in repolarization of hypocretin/orexin neurons to promote arousal. Disruption of *Kcnq2* expression during aging or experimentally through CRISPR-Cas9 drives neuronal hyperexcitability, ultimately disrupting sleep.^44^ Therefore, the sleep dysfunction seen in children with *KCNQ2*-related epilepsy may be a manifestation of a primary disease process and not necessarily secondary to seizures. Genetic differences may cause sleep dysfunction through mechanisms independent of ongoing seizure activity. We identified high rates of insomnia among individuals with *SYNGAP1*-related diagnoses, despite no documented nocturnal seizures or epileptiform activity in sleep; this is the only genetic aetiology in our cohort with complete absence of both features. Interestingly, *Syngap1* expression is regulated over the circadian cycle, with peak expression occurring during the circadian night.^42^ Further, time-of-day-gated expression of *Syngap1* is not observed in arrhythmic *Bmal1* null mice. These findings suggest that *SYNGAP1* expression may oscillate through the day and promote sleep. Melatonin showed efficacy in treating *SYNGAP1*-related sleep difficulties in prior cohorts^35^ and in our own (2/2 available reports indicated improvement).

Epileptic activity may also drive sleep dysfunction in select monogenic epilepsies. Nocturnal seizures were common among children in our cohort with *SCN1A*-related epilepsy, consistent with prior studies.^45^ In children with *SCN1A*-related Dravet Syndrome, seizures transition from predominantly prolonged, febrile seizures during wakefulness to frequent nocturnal, sleep-associated seizures after age five.^37^ In our cohort, insomnia was frequently reported among individuals with *SCN1A*-related epilepsy, and was diagnosed later than age 5, suggesting that sleep dysfunction may emerge after nocturnal seizures become more frequent. This temporal pattern points to aggressive nocturnal seizure management as a potential strategy for improving sleep quality. This is important because the presence of sleep-related diagnoses was associated with a lower frequency of eventual seizure freedom compared to the absence of such diagnoses across the full cohort. Reversing sleep impairment thus appears to be a fruitful step towards addressing seizure control, irrespective of the genetic diagnosis.

Sleep disorders were underdiagnosed within our cohort. While only 25% of individuals carried formal sleep disorder diagnoses, an additional 33% had documented sleep symptoms without formal recognition, bringing total documented sleep involvement to 58%. Insomnia was the most commonly undocumented feature, followed by sleep-disordered breathing and hypersomnolence. This diagnostic gap has clinical consequences: Sleep medications were concentrated among individuals with formal diagnoses despite substantial symptom burden in the undiagnosed group, indicating that many individuals with functionally impairing sleep difficulties are not medically managed. Given the association between sleep dysfunction and seizure persistence observed in this cohort, systemic screening for sleep difficulties may represent an underutilized opportunity to improve both sleep quality and seizure outcomes in genetically defined epilepsies.

Neuropsychiatric comorbidities, particularly ASD and ADHD, remained associated with sleep burden after adjustment for seizure severity. This underscores the importance of screening for and treating sleep difficulties in individuals with neuropsychiatric comorbidities in parallel with seizure management. *WDR45*-, *PCDH19*- and *SYNGAP1*-related neurodevelopmental disorders showed notably high rates of co-occurring ASD, ADHD and mood disorders (Table 1), with ASD present in 60-69% of individuals. This pattern has been previously appreciated for *SYNGAP1*-related diagnoses^34,35,46,47^ and extends the known phenotypic spectrum for *WDR45* and *PCDH19*. At the gene level, the co-occurrence of ASD with sleep diagnoses in *SCN2A*-related epilepsy (7/7, 100%) suggests that in certain genetic aetiologies, ASD and sleep dysfunction may be tightly linked.

Across genetic aetiologies, persistent seizures were associated with substantially greater sleep burden, with particularly strong associations observed for nocturnal and sleep-wake seizures, sleep disorder diagnoses and specific sleep subtypes including hypersomnolence and insomnia. The bidirectional relationship between sleep and epilepsy is well-established, with seizures occurring preferentially during non-rapid eye movement sleep (NREM) sleep and sleep disruption worsening seizure control.^48,49^ Sleep symptoms were associated with objective sleep architecture disruption even in the absence of formal diagnosis , indicating that patient-reported sleep difficulties correspond to measurable EEG abnormalities.

Future studies may address how different combinations of anti-seizure, psychiatric and sleep regimens alter both sleep and seizure outcomes, particularly among individuals with the monogenic diagnoses highlighted here. Anti-seizure medications can have differential effects on sleep architecture,^49^ with some agents such as lamotrigine and phenobarbital potentially worsening insomnia and daytime sleepiness, respectively, while perampanel and lacosamide may improve these symptoms, respectively.^40^ Perampanel, specifically, has shown promise in treating some monogenic epilepsies, including preclinical *SYNGAP1* models with non-REM epileptic activity.^41^ Agents that decrease NREM sleep, when seizures tend to cluster, and promote REM sleep may protect against both sleep dysfunction and nocturnal epileptic events.^42,43^ Interventions such as ketogenic diet, temporal lobe epilepsy surgery and vagus nerve stimulation devices, known for efficacy in treating refractory seizures, have been demonstrated to also increase REM sleep.^34^ Chronotherapeutic strategies warrant consideration, wherein ASMs are weighted toward increased dosing before times of day with heightened seizure risk (e.g. increasing a nighttime seizure dose to prevent seizures overnight during sleep). Nocturnal seizures significantly diminish when medications such as clobazam^44^, phenytoin and carbamazepine^45^ are dosed more heavily later in the day^46^

Our study has limitations including limited sample sizes of rare neurogenetic disorders. Some genetic diagnoses, such as *STXBP1*, are disproportionately represented in our sample, reflecting the presence of specialized clinical and research expertise for these conditions at our institution. Additionally, we described sleep/epilepsy endophenotypes by examining sleep diagnoses, sleep EEGs and self-reported sleep/wake disturbances. Polysomnograms were not available for all individuals. The presence of reported sleep symptoms does not necessarily indicate that an individual meets diagnostic criteria for a formal sleep diagnosis; accordingly, the gap between symptom prevalence and formal diagnosis rates in this cohort may reflect both true underdiagnosis and subclinical or transient sleep difficulties. Our analysis provides cross-sectional characterization rather than longitudinal tracking of symptom progression.

These findings establish sleep dysfunction as a clinically actionable yet underrecognized target in monogenic epilepsies. These endophenotypes are variable and represent an opportunity for therapeutic intervention. Sleep-related disorders beyond obstructive sleep apnoea deserve more attention to improve diagnosis and treatment.

## Data Availability

De-identified data supporting the conclusions of this study are available upon request from the corresponding author.

## Acknowledgements

This section is not mandatory.

## Funding

We thank the National Institutes of Health grant K08NS131602 and CURE Epilepsy Taking Flight Award for support to VAC. This study was made possible by the McNair Medical Institute at the Robert and Janice McNair Foundation, the Gordon and Mary Cain Research Foundation Laboratories and the Research Vision at Texas Children’s Hospital through VAC. This study was also funded by the NIH National Institute for Neurological Disorders and Stroke (R01 NS127830 and R01 NS131512, R03 HD122148, U24 NS120854 to IH)

## Competing interests

The authors report no competing interests.

## Notes

### Competing Interest Statement

The authors have declared no competing interest.

### Author Declarations

IRB of Children's Hospital of Philadelphia gave ethical approval for this work

## References

1. Fiest KM, Sauro KM, Wiebe S, et al. Prevalence and incidence of epilepsy. Neurology 2017;88:296–303.

2. Sen A, Jette N, Husain M, Sander JW. Epilepsy in older people. The Lancet 2020;395:735–748.

3. Pal DK, Pong AW, Chung WK. Genetic evaluation and counseling for epilepsy. Nature Reviews Neurology 2010 6:8 2010;6:445–453.

4. Symonds JD, Zuberi SM, Stewart K, et al. Incidence and phenotypes of childhood-onset genetic epilepsies: a prospective population-based national cohort. Brain 2019;142:2303–2318.

5. Guerrini R, Balestrini S, Wirrell EC, Walker MC. Monogenic Epilepsies: Disease Mechanisms, Clinical Phenotypes, and Targeted Therapies. Neurology 2021;97:817–831.

6. Chipaux M, Szurhaj W, Vercueil L, et al. Epilepsy diagnostic and treatment needs identified with a collaborative database involving tertiary centers in France. Epilepsia 2016;57:757–769.

7. Sultana B, Panzini MA, Veilleux Carpentier A, et al. Incidence and Prevalence of Drug-Resistant Epilepsy: A Systematic Review and Meta-analysis. Neurology 2021;96:805–817.

8. B A Wroblewski, J M Leary, A M Phelan, et al. Methylphenidate and seizure frequency in brain injured patients with seizure disorders. J. Clin. Psychiatry 1992;53:86–89.

9. Specchio LM, Iudice A, Specchio N, et al. Citalopram as treatment of depression in patients with epilepsy. Clin. Neuropharmacol. 2004;27:133–136.

10. Favale E, Audenino D, Cocito L, Albano C. The anticonvulsant effect of citalopram as an indirect evidence of serotonergic impairment in human epileptogenesis. Seizure 2003;12:316–318.

11. Brandt C, Schoendienst M, Trentowska M, et al. Efficacy and safety of pregabalin in refractory focal epilepsy with and without comorbid anxiety disorders - Results of an open-label, parallel group, investigator-initiated, proof-of-concept study. Epilepsy and Behavior 2013;29:298–304.

12. Nobili L, Beniczky S, Eriksson SH, et al. Expert Opinion: Managing sleep disturbances in people with epilepsy. Epilepsy and Behavior 2021;124

13. van Golde EGA, Gutter T, de Weerd AW. Sleep disturbances in people with epilepsy; prevalence, impact and treatment. Sleep Med. Rev. 2011;15:357–368.

14. 14. El Halal C dos S, Nunes ML. Sleep disturbances in children and adolescents with epilepsy: clinical, polysomnographic and management aspects. Sleep Med. 2026;140:108794.

15. Smith L, Singhal N, El Achkar CM, et al. PCDH19-Related Epilepsy is Associated with a Broad Neurodevelopmental Spectrum. Epilepsia 2018;59:679.

16. Buratti L, Natanti A, Viticchi G, et al. Impact of sleep disorders on the risk of seizure recurrence in juvenile myoclonic epilepsy. Epilepsy and Behavior 2018;80:21–24.

17. Kataria L, Vaughn B V. Sleep and Epilepsy. Sleep Med. Clin. 2016;11:25–38.

18. Dinopoulos A, Tsirouda MA, Bonakis A, et al. Sleep architecture and epileptic characteristics of drug naïve patients in childhood absence epilepsy spectrum. A prospective study. Seizure 2018;59:99–107.

19. Planas-Ballvé A, Grau-López L, Jiménez M, et al. El insomnio y la pobre calidad de sueño se asocian a un mal control de crisis en pacientes con epilepsia. Neurología 2022;37:639–646.

20. Lin Z, Si Q, Xiaoyi Z. Obstructive sleep apnoea in patients with epilepsy: a meta-analysis. Sleep and Breathing 2016 21:2 2016;21:263–270.

21. Pornsriniyom D, Kim H won, Bena J, et al. Effect of positive airway pressure therapy on seizure control in patients with epilepsy and obstructive sleep apnea. Epilepsy and Behavior 2014;37:270–275.

22. Elkhayat HA, Hassanein SM, Tomoum HY, et al. Melatonin and Sleep-Related Problems in Children With Intractable Epilepsy. Pediatr. Neurol. 2010;42:249–254.

23. Verma N, Maiti R, Mishra BR, et al. Effect of add-on melatonin on seizure outcome, neuronal damage, oxidative stress, and quality of life in generalized epilepsy with generalized onset motor seizures in adults: A randomized controlled trial. J. Neurosci. Res. 2021;99:1618–1631.

24. Roliz AH, Kothare S. The Interaction Between Sleep and Epilepsy. Curr. Neurol. Neurosci. Rep. 2022;22:551–563.

25. Jin B, Aung T, Geng Y, Wang S. Epilepsy and Its Interaction With Sleep and Circadian Rhythm. Front. Neurol. 2020;11:516572.

26. Yazdi Z, Sadeghniiat-Haghighi K, Naimian S, et al. Prevalence of Sleep Disorders and their Effects on Sleep Quality in Epileptic Patients. Basic Clin. Neurosci. 2013;4:36.

27. Grigg-Damberger M, Foldvary-Schaefer N. Bidirectional relationships of sleep and epilepsy in adults with epilepsy. Epilepsy and Behavior 2021;116

28. Doucoure A, Patel H, Abboud MA, et al. Biphasic sleep is a feature of sleep disturbance in SYNGAP1-RD. Sleep Med. 2026;108908.

29. Kahen A, Kavus H, Geltzeiler A, et al. Neurodevelopmental phenotypes associated with pathogenic variants in SLC6A1. J. Med. Genet. 2022;59:536–543.

30. Licheni SH, Mcmahon JM, Schneider AL, et al. Sleep problems in Dravet syndrome: a modifiable comorbidity. Dev. Med. Child Neurol. 2018;60:192–198.

31. Schoonjans AS, De Keersmaecker S, Van Bouwel M, Ceulemans B. More daytime sleepiness and worse quality of sleep in patients with Dravet Syndrome compared to other epilepsy patients. European Journal of Paediatric Neurology 2019;23:61–69.

32. Crawford K, Xian J, Helbig KL, et al. Computational analysis of 10,860 phenotypic annotations in individuals with SCN2A-related disorders. Genetics in Medicine 2021;23:1263–1272.

33. Boets S, Johannesen KM, Destree A, et al. Adult phenotype of KCNQ2 encephalopathy. J. Med. Genet. 2022;59:528–535.

34. Smith-Hicks C, Wright D, Kenny A, et al. Sleep Abnormalities in the Synaptopathies—SYNGAP1-Related Intellectual Disability and Phelan–McDermid Syndrome. Brain Sciences 2021, Vol. 11, 2021;11

35. Vlaskamp DRM, Shaw BJ, Burgess R, et al. SYNGAP1 encephalopathy: A distinctive generalized developmental and epileptic encephalopathy. Neurology 2019;92:E96–E107.

36. Sanchez REA, Bussi IL, Ben-Hamo M, et al. Circadian regulation of sleep in a pre-clinical model of Dravet syndrome: dynamics of sleep stage and siesta re-entrainment. Sleep 2019;42:1–11.

37. Han S, Yu FH, Schwartz MD, et al. Na V1.1 channels are critical for intercellular communication in the suprachiasmatic nucleus and for normal circadian rhythms. Proc. Natl. Acad. Sci. U. S. A. 2012;109:E368–E377.

38. Zhang J, Chen X, Eaton M, et al. Severe deficiency of the voltage-gated sodium channel NaV1.2 elevates neuronal excitability in adult mice. Cell Rep. 2021;36:109495.

39. Spratt PWE, Alexander RPD, Ben-Shalom R, et al. Paradoxical hyperexcitability from NaV1.2 sodium channel loss in neocortical pyramidal cells. Cell Rep. 2021;36:109483.

40. Zhang T, Yu F, Xu H, et al. Dysregulation of REV-ERBα impairs GABAergic function and promotes epileptic seizures in preclinical models. Nature Communications 2021 12:1 2021;12:1216-.

41. Wu H, Liu Y, Liu L, et al. Decreased expression of the clock gene Bmal1 is involved in the pathogenesis of temporal lobe epilepsy. Molecular Brain 2021 14:1 2021;14:113-.

42. Aten S, Kalidindi A, Yoon H, et al. SynGAP is expressed in the murine suprachiasmatic nucleus and regulates circadian-gated locomotor activity and light-entrainment capacity. European Journal of Neuroscience 2021;53:732–749.

43. Hamdy MM, Elfatatry AM, Mekky JF, Hamdy E. Rapid eye movement (REM) sleep and seizure control in idiopathic generalized epilepsy. Epilepsy and Behavior 2020;107

44. Li S Bin, Damonte VM, Chen C, et al. Hyperexcitable arousal circuits drive sleep instability during aging. Science (1979). 2022;375

45. Losito E, Kuchenbuch M, Chemaly N, et al. Age-related “Sleep/nocturnal” tonic and tonic clonic seizure clusters are underdiagnosed in patients with Dravet Syndrome. Epilepsy and Behavior 2017;74:33–40.

46. Paasch V, Doucoure A, Bifano M, Smith-Hicks CL. An exploratory study of sleep quality and quantity in children with causal variants in SYNGAP1, an autism risk gene. Sleep Med. 2023;107:101–107.[cited 2026 Mar 29 ]

47. Wright D, Kenny A, Mizen LAM, et al. Profiling Autism and Attention Deficit Hyperactivity Disorder Traits in Children with SYNGAP1-Related Intellectual Disability. Journal of Autism and Developmental Disorders 2023 55:1 2023;55:297–309.

48. Sunwoo J-S. Influence of sleep on seizures and interictal epileptiform discharges in epilepsy. Encephalitis 2024;5:1.

49. Moore JL, Carvalho DZ, St Louis EK, Bazil C. Sleep and Epilepsy: a Focused Review of Pathophysiology, Clinical Syndromes, Co-morbidities, and Therapy. Neurotherapeutics 2021;18:170.

50. Nolan KJ, Camfield CS, Camfield PR. Coping with Dravet syndrome: parental experiences with a catastrophic epilepsy. Dev. Med. Child Neurol. 2006;48:761–765.

51. Ma Z, Eaton M, Liu Y, et al. Deficiency of autism-related Scn2a gene in mice disrupts sleep patterns and circadian rhythms. Neurobiol. Dis. 2022;168:105690.

52. Hayflick SJ, Kruer MC, Gregory A, et al. Beta-propeller protein-associated neurodegeneration: a new X-linked dominant disorder with brain iron accumulation. Brain 2013;136:1708–1717.

53. Sheng D, Lv Y, Li X, et al. Case report: A young man with non-rapid eye movement parasomnias in a KCNT1-related epilepsy family. Front. Neurol. 2023;14

54. Lyu S, Xing H, Liu Y, et al. Further Studies on the Role of BTBD9 in the Cerebellum, Sleep-like Behaviors and the Restless Legs Syndrome. Neuroscience 2022;505:78–90.

55. Hackenberg A, Baumer A, Sticht H, et al. Infantile epileptic encephalopathy, transient choreoathetotic movements, and hypersomnia due to a de novo missense mutation in the scn2a gene. Neuropediatrics 2014;45:261–264.

56. Xu XH, Qiu MH, Dong H, et al. GABA transporter-1 inhibitor NO-711 alters the EEG power spectra and enhances non-rapid eye movement sleep during the active phase in mice. European Neuropsychopharmacology 2014;24:585–594.

57. Chaturvedi R, Stork T, Yuan C, et al. Astrocytic GABA transporter controls sleep by modulating GABAergic signaling in Drosophila circadian neurons. Current Biology 2022;32:1895–1908.e5.

58. Wang R, Teng S, Turanchik M, et al. Tonic-clonic seizures induce hypersomnia and suppress rapid eye movement sleep in mouse models of epilepsy. Sleep Adv. 2025;6

59. Xian J, Parthasarathy S, Ruggiero SM, et al. Assessing the landscape of STXBP1-related disorders in 534 individuals. Brain 2022;145:1668–1683.

60. Sullivan BJ, Ammanuel S, Kipnis PA, et al. Low-Dose Perampanel Rescues Cortical Gamma Dysregulation Associated With Parvalbumin Interneuron GluA2 Upregulation in Epileptic Syngap1+/− Mice. Biol. Psychiatry 2020;87:829–842.

61. Dhamija R, Erickson MK, St Louis EK, et al. Sleep Abnormalities in Children With Dravet Syndrome. Pediatr. Neurol. 2014;50:474–478.

62. Papale LA, Makinson CD, Christopher Ehlen J, et al. Altered Sleep Regulation in a Mouse Model of SCN1A-Derived Genetic Epilepsy with Febrile Seizures Plus (GEFS+). Epilepsia 2013;54:625.

63. Kalume F, Oakley JC, Westenbroek RE, et al. Sleep impairment and reduced interneuron excitability in a mouse model of Dravet Syndrome. Neurobiol. Dis. 2015;77:141–154.

64. Xu XH, Qu WM, Bian MJ, et al. Essential Roles of GABA Transporter-1 in Controlling Rapid Eye Movement Sleep and in Increased Slow Wave Activity after Sleep Deprivation. PLoS One 2013;8:e75823.

65. Lamers D, Landi S, Mezzena R, et al. Perturbation of Cortical Excitability in a Conditional Model of PCDH19 Disorder. Cells 2022, Vol. 11, 2022;11

66. Milh M, Villeneuve N, Chouchane M, et al. Epileptic and nonepileptic features in patients with early onset epileptic encephalopathy and STXBP1 mutations. Epilepsia 2011;52:1828–1834.

67. Wilson JL, Gregory A, Kurian MA, et al. Consensus clinical management guideline for beta-propeller protein-associated neurodegeneration. Dev. Med. Child Neurol. 2021;63:1402–1409.

68. Kidokoro H, Yamamoto H, Kubota T, et al. High-amplitude fast activity in EEG: An early diagnostic marker in children with beta-propeller protein-associated neurodegeneration (BPAN). Clinical Neurophysiology 2020;131:2100–2104.

69. Miceli F, Millevert C, Soldovieri MV, et al. KCNQ2 R144 variants cause neurodevelopmental disability with language impairment and autistic features without neonatal seizures through a gain-of-function mechanism. EBioMedicine 2022;81:104130.

70. Ikeda H, Imai K, Ikeda H, et al. Characteristic phasic evolution of convulsive seizure in PCDH19-related epilepsy. Epileptic Disorders 2016;18:26–33.

71. Creson TK, Rojas C, Hwaun E, et al. Re-expression of SynGAP protein in adulthood improves translatable measures of brain function and behavior. Elife 2019;8[cited 2026 Mar 29 ]

72. Gerbatin RR, Augusto J, Boutouil H, et al. Life-span characterization of epilepsy and comorbidities in Dravet syndrome mice carrying a targeted deletion of exon 1 of the Scn1a gene. Exp. Neurol. 2022;354:114090.

73. Kessi M, Peng J, Yang L, et al. Genetic etiologies of the electrical status epilepticus during slow wave sleep: systematic review. BMC Genetics 2018 19:1 2018;19:40-.

74. Miao P, Tang S, Ye J, et al. Differential Functional Changes of Nav1.2 Channel Causing SCN2A-Related Epilepsy and Status Epilepticus During Slow Sleep. Front. Neurol. 2021;12:653517.

75. Johannesen KM, Gardella E, Linnankivi T, et al. Defining the phenotypic spectrum of SLC6A1 mutations. Epilepsia 2018;59:389–402.

76. Boillot M, Baulac S. Genetic models of focal epilepsies. J. Neurosci. Methods 2016;260:132–143.

77. Cataldi M, Nobili L, Zara F, et al. Migrating focal seizures in Autosomal Dominant Sleep-related Hypermotor Epilepsy with KCNT1 mutation. Seizure 2019;67:57–60.

78. Heron SE, Smith KR, Bahlo M, et al. Missense mutations in the sodium-gated potassium channel gene KCNT1 cause severe autosomal dominant nocturnal frontal lobe epilepsy. Nature Genetics 2012 44:11 2012;44:1188–1190.

79. Stamberger H, Nikanorova M, Willemsen MH, et al. STXBP1 encephalopathy. Neurology 2016;86:954–962.

80. Huang CH, Hung PL, Fan PC, et al. Clinical spectrum and the comorbidities of Dravet syndrome in Taiwan and the possible molecular mechanisms. Scientific Reports 2021 11:1 2021;11:20242-.

81. Yuan F fen, Gu X, Huang X, et al. SLC6A1 gene involvement in susceptibility to attention-deficit/hyperactivity disorder: A case-control study and gene-environment interaction. Prog. Neuropsychopharmacol. Biol. Psychiatry 2017;77:202–208.

